# Matrix and Graphical Representation of the Primary Headache Syndromes in the International Classification of Headache Disorders (ICHD3): A Basis for Automated Diagnosis and Analysis of Criteria

**DOI:** 10.64898/2026.02.02.26345241

**Authors:** Pengfei Zhang, Roger Cheng

**Author notes:** **Conflict of Interest Statement:** PZ: He has received honorarium from Acumen LLC. **Financial Support**: None. **Trial Registration**: Not applicable.

## Abstract

**Objective:** We represent primary headaches of the ICHD3 in matrix form and show that this representation allows for automated diagnosis as well as additional insights into headache classification.

**Methods:** Each diagnosis in the ICHD3 is defined by a list of characteristics; combinations of characteristics form phenotypes. Multiple phenotypes may fit a given diagnosis. We first translated all characteristics for primary headache diagnoses in the ICHD3 into true/false statements. We generated a matrix of valid ICHD3 diagnosis as follows:

1. Each row of the matrix represents a phenotype.
2. Each column of the matrix represents a characteristic.
3. If any phenotype contains a characteristic, then that element is encoded as 1. Otherwise, it is encoded as 0.

From this matrix, we calculated its bipartite projection and Markov cluster. We also row reduced to derive the basis vectors that span the space of all headache phenotypes.

**Results:** Chronic migraine diagnoses as well as the characteristics “greater than 15 days per month” and “more than 3 months” have the strongest associations based on bipartite projection. Markov clustering yields 64 clusters. These clusters can be organized by ICHD3 diagnoses and demonstrates the level of fragmentation of individual diagnosis in the classification: Migraine is composed of 1 cluster, for example, whereas paroxysmal hemicrania can be broken down into 9 clusters. Finally, row reduction of our matrix yields 63 basis vectors, implying that all headache diagnoses in the ICHD3 can be represented as linear combinations of 63 characteristics. These 63 characteristics corresponds to the following: duration, frequency, aura characteristics, size/location, laterality, clearly remembered onset, TAC features, total number of episodes, severity, nausea/vomiting, photophobia, pulsating, alleviation by triptans, and association with awakening, sexual activity, physical activity, temperature, compression or traction, coughing.

**Conclusion:** Our result demonstrates that ICHD3 is a mathematical entity and that headache diagnoses exist in a 63-dimensional vector space. This mathematical embodiment of classification allows us to conduct 1) large scale systematic investigations of relationships between headache and phenotypes, 2) generate a graphical representation of characteristics and phenotypes and 3) improves diagnostic accuracy and efficiency.

## Introduction

The International Classification of Headache Disorders (ICHD3) has been widely used as the standard for headache diagnosis in both clinical and research settings.^1^ Constructed with criteria-based language, the classification offers precise definitions for each primary and secondary headaches disorders. As a result, ICHD3 contains a logical structure.^2,3^ This paper attempts to study the logical nature of diagnostic criteria at scale. Specifically, we propose a numerical embodiment of primary headache diagnosis in ICHD3 in the form of a matrix equivalent. Since there is a natural relationship between matrices and networks, we also developed a graphical representation of the classification. We then demonstrate that this matrix/graphical form of the ICHD3 allows us to study how headache phenotypes or diagnoses are correlated due to the classification’s inherent structure. We will also show that this matrix formulation allows us to interpret diagnostic criteria, and therefore headaches in general, as members within a mathematical vector space. Finally, our matrix formulation of the international criteria allows us to automate ICHD3 diagnosis of headaches.

## Methods

We define a headache *characteristic* as a variable that takes on a Boolean value (either True or False). We define a headache *phenotype* as a combination of headache characteristics. A *headache disorder,* or simply *disorder* in this paper, is composed of one or more phenotypes. For example, “photophobia” is a headache *characteristic*. A patient with a combination of *characteristics* including headaches that are “unilateral”, “moderate to severe”, “lasting 4 hours to 3 days”, with “photophobia”, “phonophobia”, but “no nausea”, has one of the many headache *phenotypes* which satisfies criteria for a *headache disorder* called migraine without aura. (We take these definitions from prior modeling of ICHD3.^3,14^)

## Inclusion/Exclusion

In our study, we extracted the criteria for diagnosis of primary headache disorders up to two levels deep in ICHD3. We excluded “Complications of migraine” (1.5) and “Episodic syndrome that may be associated with migraine” (1.6) since these diagnoses depend on a primary diagnosis of migraine. We have retained the criteria for chronic migraine in our study due to its relevance in our field.

## Matrix construction

We constructed a matrix with the following rules:

1. We arranged phenotypes along the row of the matrix. Phenotypes are named by their ICHD3 diagnosis, followed by a numerical designation.
2. We arranged characteristics along the columns of the matrix
3. If a phenotype on row X contains a specific characteristic on column Y, then the cell (X,Y) is labeled as 1. Otherwise, it is labeled as 0.

This matrix is included as Supplement Material 1 of this paper.

## Methods of Analysis

Matrix interpretation of ICHD3 allows for numerical analysis of the relationship between phenotypes (the row) and characteristics (the column). We applied two commonly used data science techniques to our matrix: 1) bipartite projection and 2) Markov clustering. We also applied Gaussian elimination to our matrix to obtain the basis vectors in row reduced echelon form (RREF). We will provide a brief overview of each of the above methods for the lay reader; our citations provide more in-depth references for each of the methods:

A bipartite graph is a mathematical graph containing two disjoint groups of vertices where no two vertices in the same group are connected.^4^ In graphs with this kind of property, one can calculate the graph’s “bipartite projection”, a measurement of how individual vertices in one group are associated with each other based on connections to the vertices in the other group (e.g. shared neighbors).^5,6,7^ In our case, headache phenotypes and headache characteristics are two disjoint groups of nodes. An edge exists between a phenotype and a characteristic if that element of the matrix is denoted as “1”. Using bipartite projection, we can determine which phenotypes (i.e. diagnoses) are associated with each other; the strength of these associations (weight) is based on the number of shared connections to corresponding headache characteristics (the other group). The converse can also be done – associating characteristics based on connections to phenotypes. In this study, we identify the pairings of phenotypes as well as pairings of characteristics with the two highest weightings.^6^

Since the ICHD3 matrix is the adjacency matrix of a mathematical graph (Figure 1 to 6), we then apply the Markov clustering algorithm to the matrix to identify groupings within the ICHD3.^9,10^ Conceptually, Markov clustering is used to identify probability distributions that would result from random walks over a given graph (stochastic flow). Markov clustering relies on two parameters: expansion and inflation. To assess optimal parameters, we updated and applied the modularity method in Guy Allard’s Markov Clustering Python library for our purpose.^10, 11^ We then implemented the Markov clustering algorithm using custom Python code with those optimal parameters.

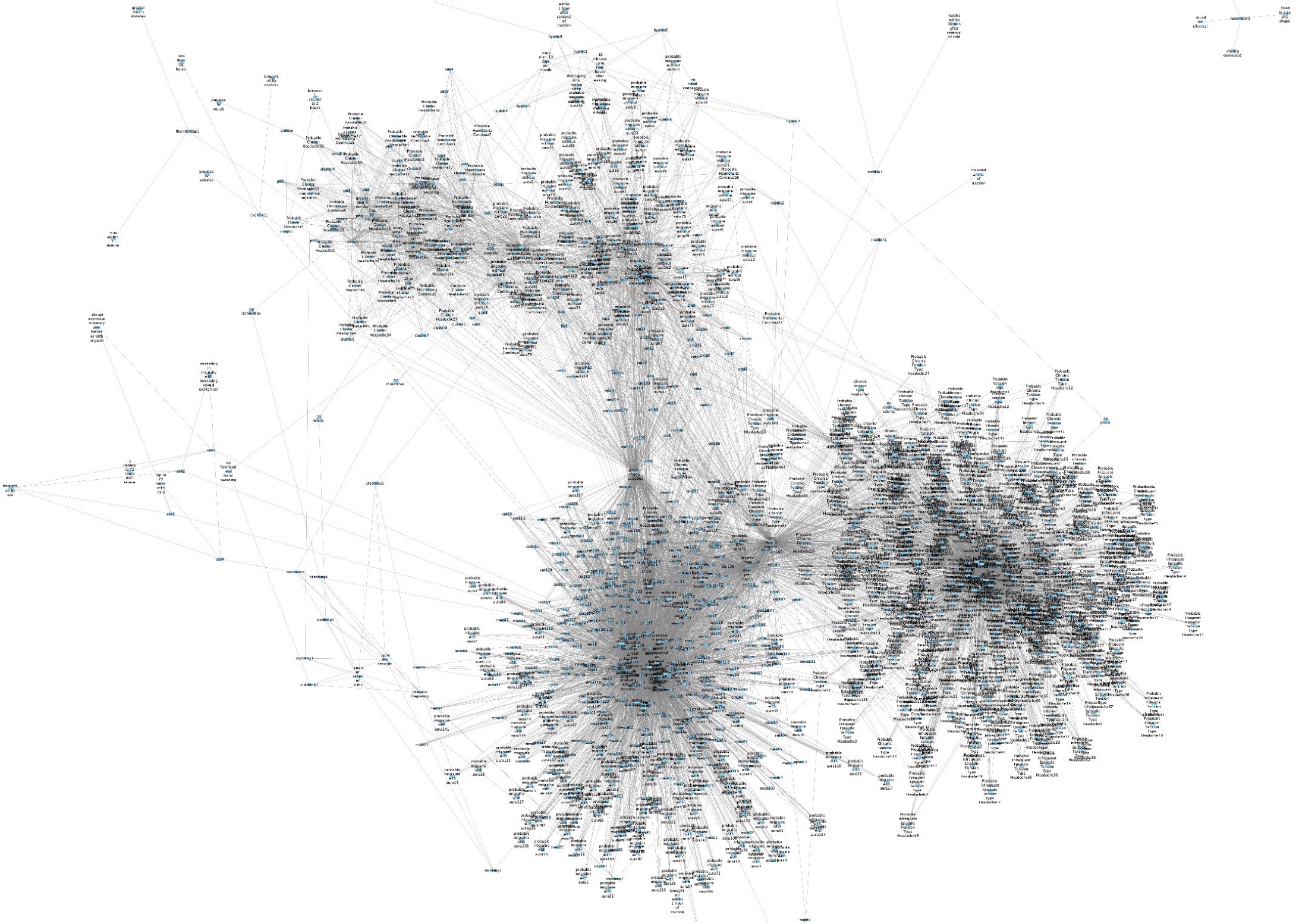

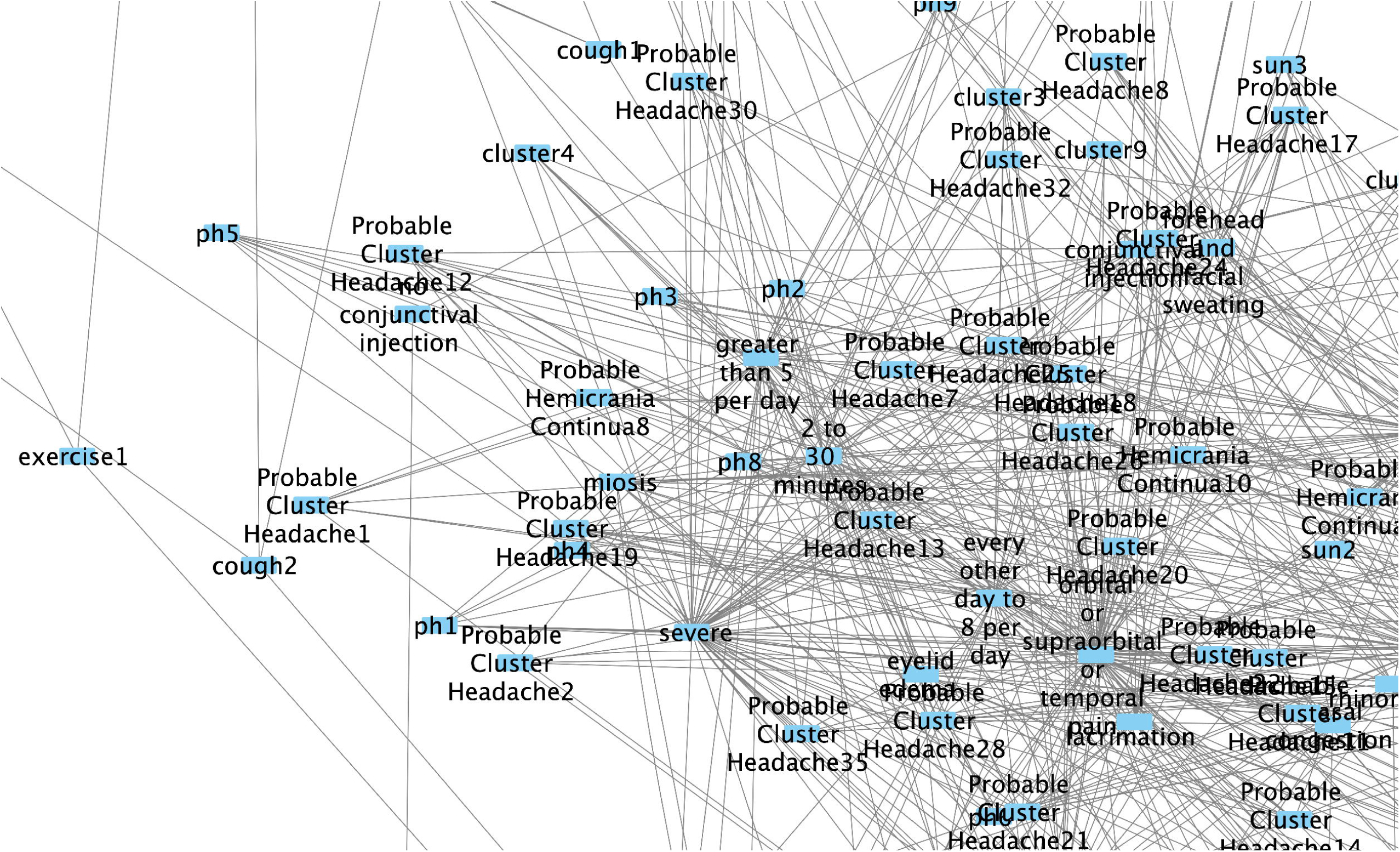

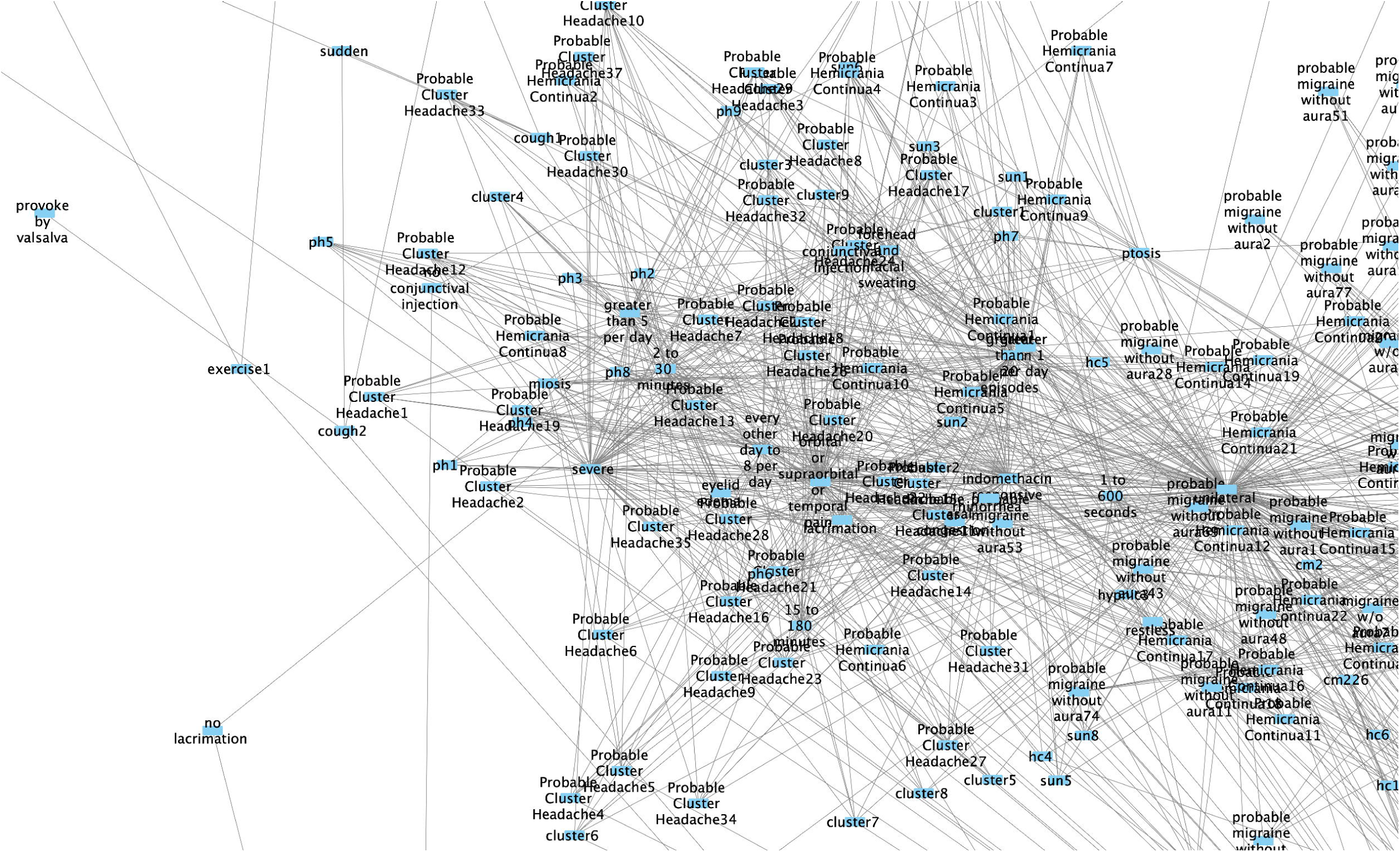

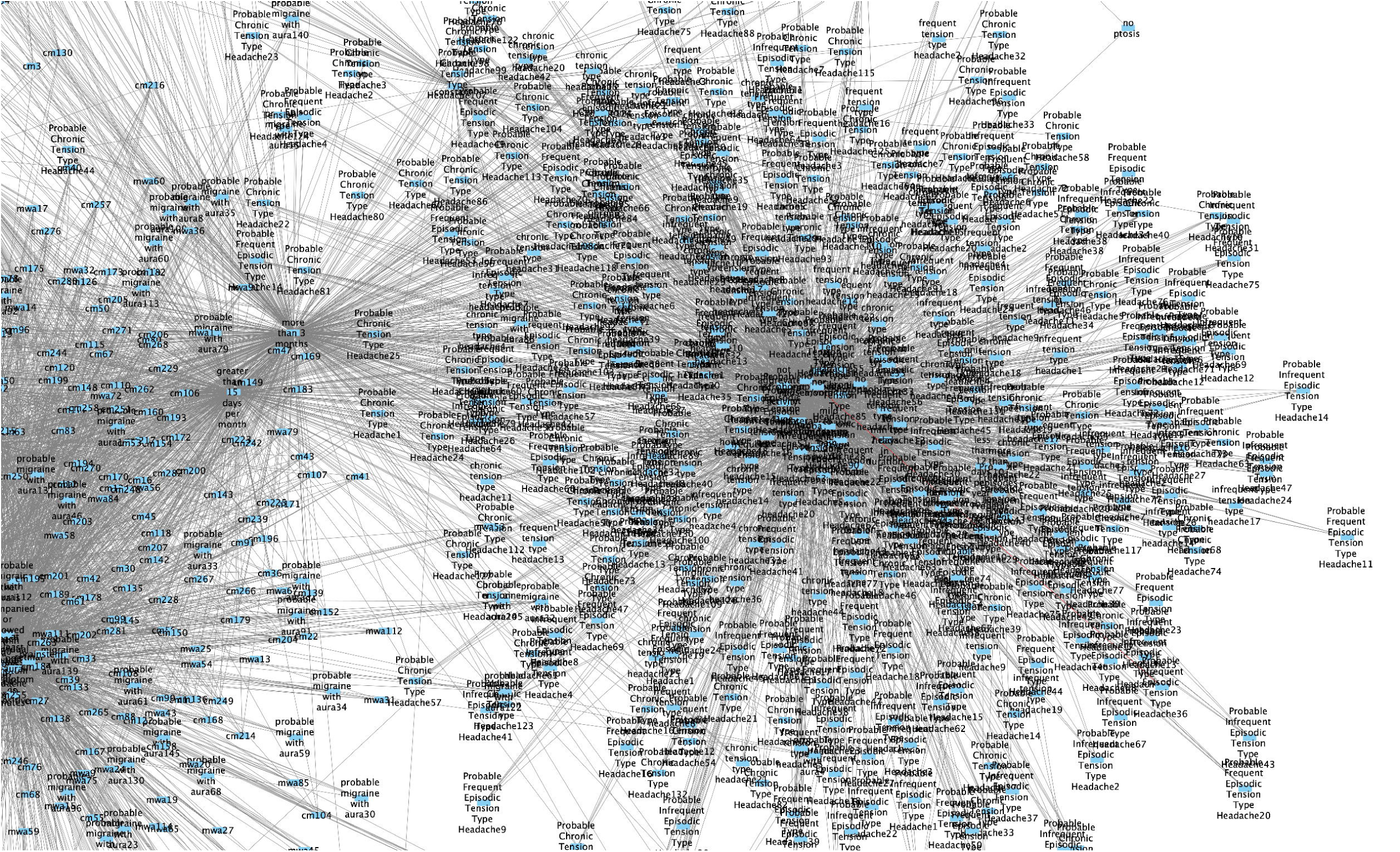

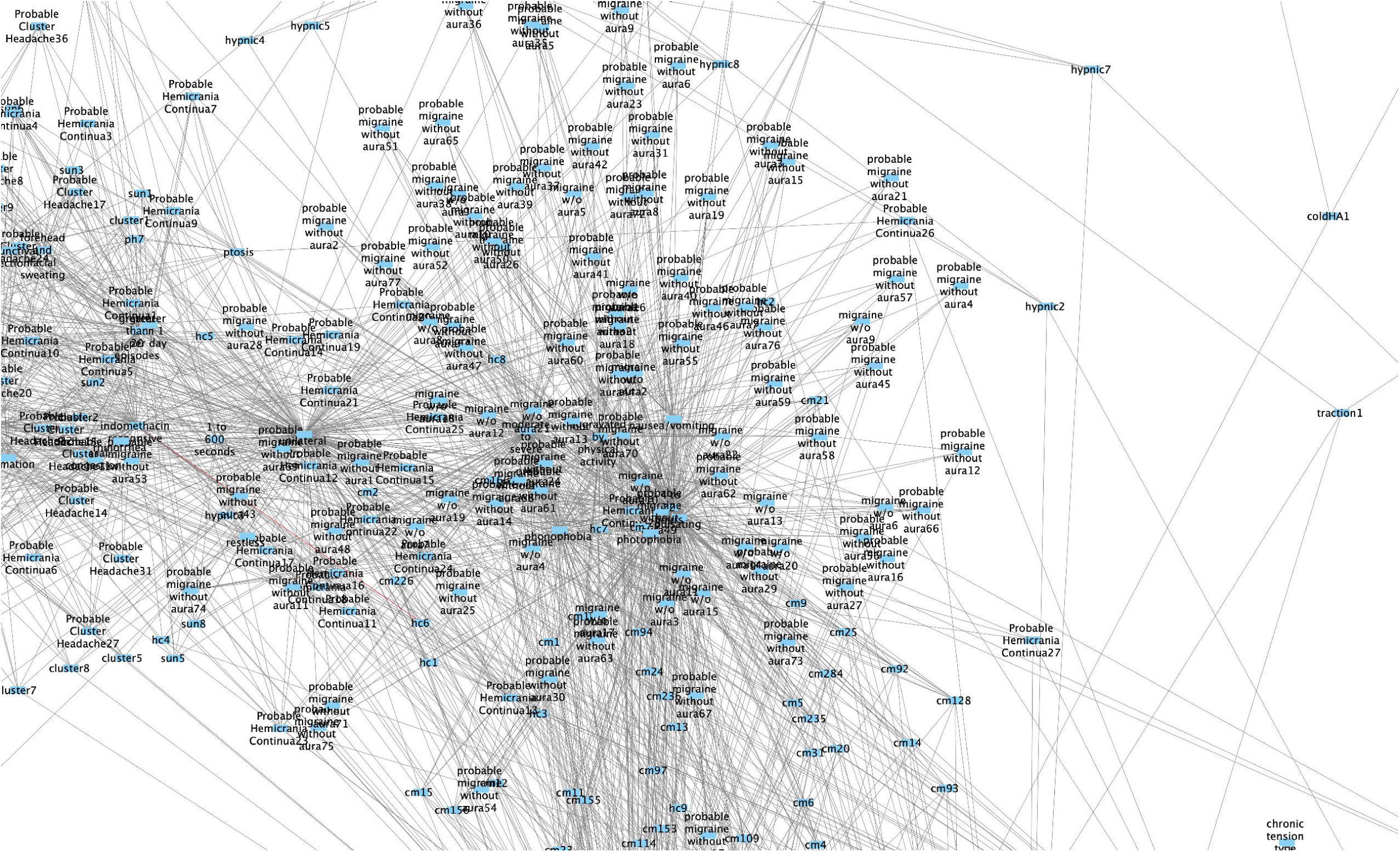

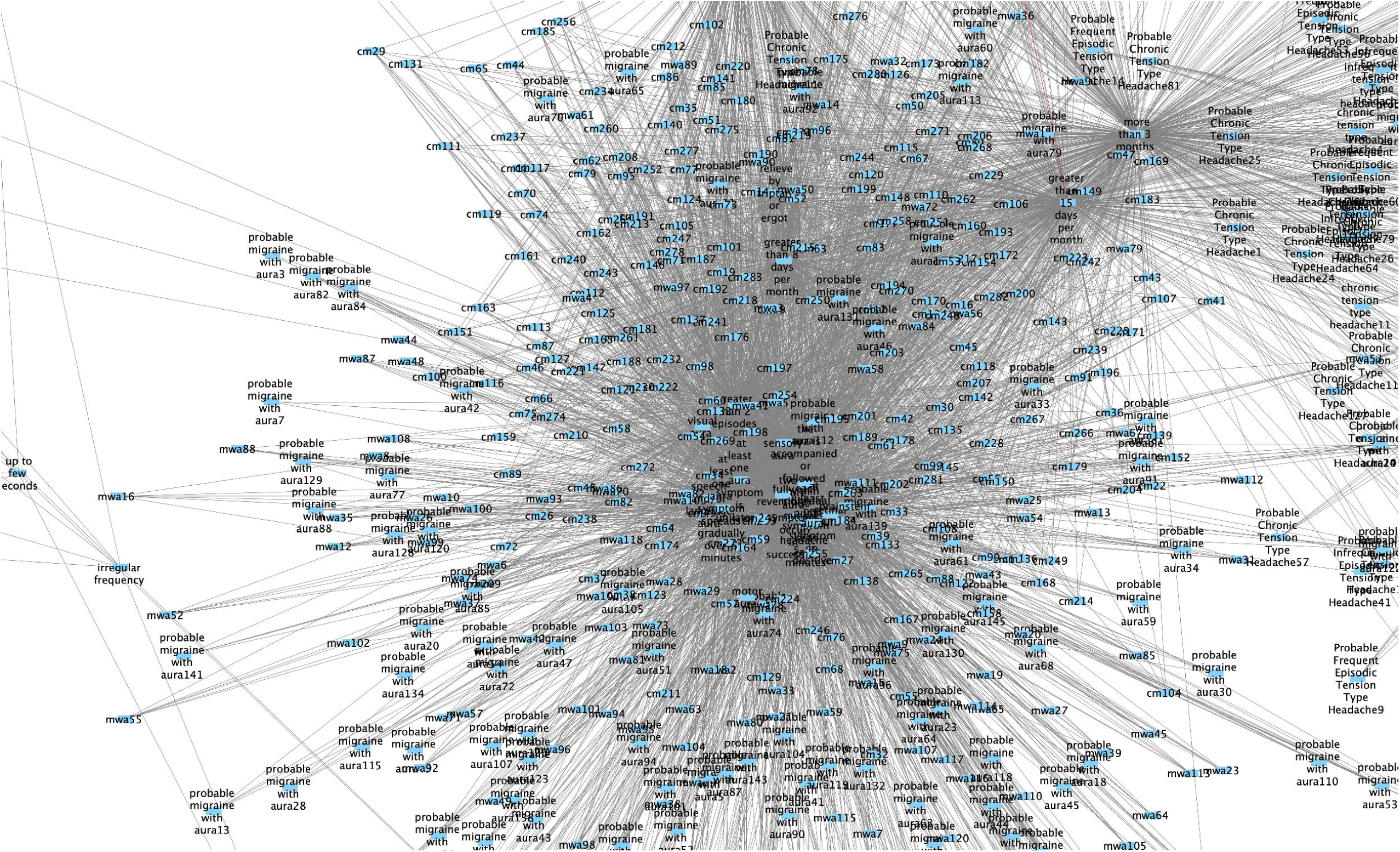

## The Vector Space of Headache Diagnosis

Algebraically, any matrix can be interpreted as a combination of vectors. The technique of Gaussian elimination - transforming matrices into row reduced echelon form (RREF) - is one way of identifying the minimum number of vectors needed to represent the same information presented in the matrix. These vectors are called basis vectors, and their possible combinations, through addition and multiplication, form a *vector space*.^15^ Our matrix is no different: the ICHD3 (our matrix) can be viewed as a combination of characteristics (vectors). We therefore applied RREF to our matrix to identify the minimum number of characteristics, which when used in combination, can describe all diagnoses in ICHD3.

### Automated Diagnosis

Finally, the matrix representation of ICHD3 can be used for automating diagnosis of headache disorders. One can reformulate a given patient’s response (0 for false and 1 for true) to each of the characteristics presented in the matrix as a vector. Multiplication of this vector with our matrix yields a new matrix, such that, if there is a row which is unchanged from the original, then that row is the diagnosis. We provide a mathematical proof of this in our supplemental material. (This proof was first presented at the 18^th^ European Headache Congress under the poster entitled “*Which types of headaches in the migraine and tension-type spectrum cannot be diagnosed by ICHD3?*” and is reproduced here with permission.)^18^ Of note, this diagnostic technique of matrix representation is equivalent to the prime number encoding which was presented in a prior project.^3^

## Implementation

The generation and extraction of ICHD3 into a matrix form is done manually and through custom code using the programming language Haskell. Once the matrix is generated, we used the Python libraries *pandas*, *matplotlib*, and *numpy* to solve for the bipartite projection. Markov clustering was accomplished with custom code as well as Allard’s *Markov Clustering* Python library.^10^ Since any adjacent matrix is a mathematical graph, we used custom Python code to translate the matrix into pairings of edges so that *Cytoscape*, version 3.10.4, a graphic/network visualization software package, can be used for graphic visualizations.^12^

## Results

Figure 1 represents a graphical representation of the ICHD3 primary headache disorders in this manuscript. To offer better visualization of the individual components, we offer a “zoomed in” figure of individual major clusters in Figure 2 to 6.

### Results from Bipartite Projection

As discussed above, bipartite projections can be done for either phenotypes or characteristics. The highest weight for phenotypes is 11; these phenotypes all consist of pairings of chronic migraine: (’cm227’, ‘cm284’), (‘cm235’, ‘cm284’), (‘cm236’, ‘cm284’), (‘cm259’, ‘cm284’), (‘cm279’, ‘cm284’). The second highest weight is 10; there are 1006 pairings, all between various phenotypes of chronic migraine as well.

The highest weight for characteristics is 416; they consist of the pairings between “greater than 15 days per month” and “more than 3 months”. The second highest weight for characteristics is 366 and is between “fully reversible” and “greater than 2 episodes”. We have also obtained the third largest weight for characteristics -284 - post hoc; the weight belongs to the following pairings: (‘greater than 15 days per month’, ‘greater than 5 episodes’), (‘greater than 15 days per month’, ‘greater than 8 days per month’), (‘greater than 5 episodes’, ‘greater than 8 days per month’), (‘greater than 5 episodes’, ‘more than 3 months’), and (‘greater than 8 days per month’, ‘more than 3 months’).

### Results from Markov clustering

Since inflation and expansion parameters may take on any values, modularity measurement is a method deployed to determine the optimal parameter values in Markov clustering.^11^ Using modularity measurement, the inflation parameter is optimal at 3.3. Given this inflation parameter, expansion is then determined - through modularity measurement again - as optimal at 3 (See Supplementary Materials under “modularityTestingOnExpansion.txt” and “modularityTestingResultOnInflation.txt”) Applications of these parameters yields 64 clusters. (See Supplementary Materials. “totalClusterApp.txt”) We manually reviewed each of these clusters and categorized them into those dominated by specific ICHD3 diagnoses. They are as follows:

1. Trigeminal autonomic cephalalgias (TAC)

a. Clusters 8 to 16 (paroxysmal hemicrania only)
b. Cluster 17 (SUN, Probable HC, “greater than 1 per day”, “1 to 600 seconds”)
c. Cluster 7 (cluster headache)
d. Cluster 3 (cluster headache, probable cluster headaches, thunderclap headaches, “maximum within 1 min”, “greater than 5 minutes”)
e. Cluster 22 (Probable Cluster Headache, Probable Hemicrania Continua, “greater 5 per day”, “miosis”)
f. Cluster 23 (Probable Hemicrania Continua, Hemicrania Continua, Probable Cluster Headache, “greater than 20 episodes”, “nasal congestion”)
g. Cluster 20 (Probable Hemicrania Continua, Hemicrania Continua, Probable Cluster Headache, “forehead and facial sweating”, “indomethacin responsive”)
h. Cluster 21 (Probable Hemicrania Continua13, Probable Cluster Headache4, hc4, “orbital or supraorbital or temporal pain”, “lacrimation”)
i. Cluster 25 (Probable Cluster Headache, Probable Hemicrania Continua, severe, every other day to 8 per day, 15 to 180 minutes, restless)
j. Cluster 19 (Probable Cluster Headache, Probable Hemicrania Continua, hc2, unilateral, eyelid edema)
k. Cluster 18 (Probable Cluster Headache1, Probable Hemicrania Continua15, hc1, 2 to 30 minutes, conjunctival injection)
l. Cluster 26 (Probable Cluster Headache, hc9, Probable Hemicrania Continua25, rhinorrhea)
m. Cluster 24 (Probable Hemicrania Continua19, Probable Cluster Headache7, hc7, ptosis)
2. Tension type headaches (TTH)

a. Cluster 1 (NDPH, frequent TTH, chronic TTH, probable chronic TTH, probable frequent TTH, “constant”, “not aggravated by activity”, “mild to moderate pain”, “no photophobia”, “clearly remembered onset”, “no phonophobia”, “no nausea/vomiting”, “30 min to 7 days in duration”, “1 to 14 days per month”, “more than 3 months”, “non-pulsating”, “unremitting within 24 hours”, “hours to days”, “bilateral location”)
b. Cluster 39 to 60 (infrequent tension type headaches only)
c. Cluster 61 (probable infrequent tension type headaches, “less than 12 days per year”, “more than 10 episodes”)
d. Cluster 34 (Probable Chronic Tension Type Headache, stabbing, hypnic, no ptosis)
3. Migraine (with and without aura):

a. Cluster 0 (chronic migraine, migraine with aura, migraine without aura, probable migraine with aura, probable migraine without aura, “relieve by triptans or ergots”, “moderate to severe”, “at least one aura symptom is unilateral”, “visual aura”, “greater than 15 days per month”, “fully reversible”, “aggravated by physical activities”, “photophobia”, “motor aura”, “phonophobia”, “aura is accompanied, or followed within 60 min by headaches”, “two or more aura symptoms occur in succession”, “pulsating”, “sensory aura”, “at least one aura symptom is positive”, “greater than 2 episodes”, “speech and/or language aura”, “nausea/vomiting”, “4 to 72 hours”, “each individual aura symptom lasts 5 to 60 minutes”, “greater than 5 episodes”, “at least one aura symptom spreads gradually over 5 minutes”, “brainstem aura”, “greater than 8 days per month”, “retinal aura”)
4. Primary sex headache:

a. Cluster 27 (probable migraine with aura, sex, “abrupt explosive intensity just before or with orgasm”, “1 minute to 72 hours with severe”, “up to 72 hours with mild”, “increasing in intensity with increasing sexual excitement”, “brought on by sex”)
5. Primary stabbing and hypnic headache:

a. Cluster 28 (primary stabbing headaches, hypnic headaches, “15 minutes up to four hours after waking”, “more than 10 days per month”, “developing only during sleep and causing wakening”, “no conjunctival injection”)
b. Cluster 29 (primary stabbing headaches, hypnic headaches, “no eyelid edema”)
c. Cluster 35 (hypnic, primary stabbing, “no rhinorrhea”)
d. Cluster 31 (stabbing4, Probable Chronic Tension Type Headache23, hypnic4, “no lacrimation”)
e. Cluster 30 (hypnic3, stabbing3, Probable Chronic Tension Type Headache20, no forehead and facial sweating)
f. Cluster 32 (stabbing5, hypnic5, single or series of stabs, no miosis, irregular frequency, up to few seconds)
g. Cluster 33 (hypnic6, stabbing6, “no nasal congestion”)
6. Cough and Valsalva headache:

a. Cluster 37(cough1, “sudden”, “between 1 second to 2 hours”, “provoke by cough”)
b. Cluster 38(cough headache, “provoke by Valsalva”)
7. Exercise headaches:

a. Cluster 2 (exercise headaches, brough on by exercise, less than 48 hours)
8. Traction and compression headaches:

a. Cluster 5 (compression headaches, “brought on within 1 hour of compression”, “maximal at site of compression”, “resolve within 1 hour after removal of compression”)
b. Cluster 6 (traction headaches, “brought on within 1 hour of traction”, “maximal at site of traction”, “resolve within 1 hour after removal of traction”
9. Cold induced headaches:

a. Cluster 4 (cold induced HA, “brought on by cold stimuli”, “resolve within 30 min after removal of cold”)
10. Nummular headache:

a. Cluster 36 (nummular1, fixed in size and shape, sharply contoured, 1–6 cm in diameter, round or elliptical)
11. Phenotypes only:

a. Cluster 62 (more than 1 episode per day)
b. Cluster 63 (no orbital or supraorbital or temporal pain)
c. Cluster 64 (no restless)

### Row reduction

The pivot columns/basis vectors for all diagnoses (excluding probable diagnoses) are the following 63 characteristics: “1 minute to 72 hours with severe pain”, “1 to 14 days per month”, “1 to 600 seconds”, “15 minutes to up to four hours after waking”, “15 to 180 minutes”, “1 to 6cm in diameter”, “2 to 30 minutes”, “30 minutes to 7 days in duration”, “4 to 72 hours”, “abrupt explosive intensity just before or with orgasm”, “aggravated by physical activity”, “at least one aura symptom is positive”, “at least one symptom is unilateral”, “at least one symptom spreads gradually”, “between 1 second to 2 hours”, “bilateral location”, “brainstem aura”, “brough on by cold stimuli”, “brough on by exercise”, “brought on by sex”, “brought on within 1 hour of compression”, “brought on within 1 hour of traction”, “clearly remembered onset”, “conjunctival injection”, “constant”, “aura lasting 5 to 60 minutes”, “eyelid edema”, “forehead and facial sweating”, “fully reversible”, “greater than 15 days per month”, “greater than 5 episodes”, “greater than 5 minutes”, “greater than 8 days per month”, “hours to days”, “irregular frequency”, “lacrimation”, “mild to moderate”, “miosis”, “moderate to severe”, “motor aura”, “nasal congestion”, “nausea/vomiting”, “no conjunctival injection”, “no eyelid edema”, “no forehead and facial sweating”, “no lacrimation”, “no miosis”, “no nasal congestion”, “no phonophobia”, “no ptosis”, “no restlessness”, “non-pulsating”, “not aggravated by physical activity”, “provoked by coughing”, “ptosis”, “pulsating”, “relieved by triptan”, “restlessness”, “retinal aura”, “sensory aura”, “speech and/or language aura”, “aura is accompanied or followed within 60 minutes by headache”, and “unilateral headache”.

This corresponds to characteristics describing the following:

1. Duration
2. Frequency
3. Association with awakening
4. Association with sexual activity
5. Association with physical activity
6. Association with temperature
7. Association with compression or traction
8. Association with cough
9. Aura characterization
10. Size/location
11. Laterality
12. Clearly remembered onset
13. Existence of trigeminal autonomic features
14. Total number of episodes
15. Severity
16. Nausea/vomiting
17. Photophobia
18. Pulsating
19. Alleviation by triptan

## Discussion

In this paper, we demonstrate that the international classification can be translated into matrix form, allowing for large-scale numerical manipulations and investigation. This numerical version of our classification guidelines enables us to study the ICHD3 in the following fashion: 1) Identification of correlations between phenotypes, characteristics, and headache disorders. 2) Identification of a headache vector space as defined by characteristics. 3) The generation of automated diagnosis at scale through the use of linear algebra. 4) Production of a graphical version of the criteria. In summary, the current method shows that classification is a mathematical entity.

We should note that this method is not the only way to produce a mathematical version of ICHD3. Previous work by Costabile et al. show that ICHD3 can be interpreted logically.^2^ Our own works showed that it is possible to map ICHD3 into a number system through a version of Godel numbering.^3,14^ Furthermore, this method is also not the only way to describe ICHD3 graphically: the hierarchical nature of ICHD3 as well as its comment section allows for graphical representation of both primary and secondary headaches, which then in turn can be represented as set/subset relationships between differential diagnoses.^12,16^

The study of the structure of headache classification, although theoretical, is in reality immediately relevant to clinicians and researchers. ICHD3 guides diagnosis and research; any inherent structural bias directly influences patient care. Study of the classification is the study of our contemporary perspective of headache disorders. Our goal in this study is not only to put forth a method of studying classification as well as a method of automated diagnosis but also to point out potential diagnostic biases – both desirable and undesirable - in primary headache diagnosis.

First, the overabundance of chronic migraine in the bipartite projection highlights the importance that the various phenotypes of chronic migraine play structurally in our classification. This also implies that chronic migraine is formed by two tightly clustered and massive groups, as evident in Figure 1. These two groups are held together by two characteristics – “greater than 5 episodes” and “greater than 8 days per month”. This likely explains why various characteristics of chronic migraine also cluster together in the bipartite projection of characteristics: cf “greater than 15 days per month” and “more than 3 months”, “greater than 15 days per month” and “greater than 8 days per month”, “greater than 15 days per month” and “greater than 5 episodes”, “greater than 5 episodes” and “greater than 8 days per month”, “greater than 5 episodes” and “more than 3 months”, as well as “greater than 8 days per month” and “more than 3 months”.

Even though photophobia, phonophobia, and nausea/vomiting are also major characteristics that form the chronic migraine/migraine cluster, it is duration and frequency clusters that exert a stronger relationship. This leads to the question of whether these two characteristics should hold such a powerful role in defining chronic migraine. Although at first glance it may appear surprising why “more than 8 days per month” is an equally relevant characteristic as “more than 15 days per month”, a study of the chronic migraine criteria in the ICHD3 makes this clear – whereas “more than 15 days per month” allows for unification of migraine diagnoses under chronic migraine, “more than 8 days per month” actually allows headaches with tension type phenotype to also be grouped under the classification of “migraine”. In other words, the way the “8 days” rule in the ICHD3 - “*On 8 days/month for >3 months, fulfilling any of the following 2: 1. criteria C and D for 1.1 Migraine without aura 2. criteria B and C for 1.2 Migraine with aura 3. believed by the patient to be migraine at onset and relieved by a triptan or ergot derivative*” – allows the rest of the headache days to be counted towards migraine even if they are not “migrainous” may be a significant factor in the diagnosis of (or at least our perception of) how we should diagnose migraine.

Although characteristics of migraine with aura are unsurprisingly strongly associated – for example “fully reversible” and “greater than 2 episodes” is present as a highly correlated pairing – our study also highlights that the cardinal features of aura are reversibility and reproducibility as opposed to the specific forms of aura present. We believe it is an open question of whether this is desirable.

Markov clustering can be thought of as random walks on Figure 1. Firstly, we should make explicit our assumption of classifying Markov clusters with respect to primary headache diagnoses/phenotypes rather than the characteristics. This is an editorial decision on our part, and it may be equally valid to attempt to classify clusters with characteristics instead. However, we deem our approach more appropriate given that ICHD3 is intended to describe coherent clinically observable entities – i.e. migraines – rather than inspecting how each characteristic can be described by its disease process. So therefore, we preference grouping of phenotypes over grouping with respect to characteristics.

An obvious question is “Why are some diagnoses compose of a few big clusters but others a variety of small clusters?” For example, paroxysmal hemicrania (PH) is broken up into 9 small clusters (cluster 8 to 16) whereas migraine can be described as simply 1 big cluster (cluster 0). This fragmentation also occurs for infrequent tension type headache, which occupied 22 cluster groups (cluster 39 to 60). An explanation can be gained by looking into the mathematical graph generated by the adjacency matrix. (Figure 1). Whereas the characteristics of “more than 3 months”, “nausea/vomiting”, “greater than 5 episodes”, and other migrainous characteristics appears to have connected 2 major clusters of migraine together like spokes to a wheel, paroxysmal hemicrania of different phenotypes do not appear to share a common characteristic. (Figure 2) Rather, the shared characteristics of TACs are a set of characteristics – rhinorrhea, miosis, etc. Since any one of those characteristics is sufficient to identify a phenotype as a TAC, the graphical properties will be more divergent compared to a phenotype where one since mandatory characteristics – such as “more than 3 months” – is required for diagnosis. (Indeed, given the result from the bipartite projection – which highlights to close correlation between various phenotypes of chronic migraine – there is no surprise that migraine subtypes are so closely intertwined - as opposed to tension type headache - for example.)

In other words, the logical AND statement in the criteria is more unifying than OR statements. In some sense, this should not be surprising: after all, there must be some fundamental pathophysiological differences between a PH patient with rhinorrhea as compared to one with miosis, even if they are both classifiable under the same umbrella condition.

Paroxysmal hemicrania appears to exist in its own distinct clusters. (Cluster 8 to 16) The same can be said of short-acting unilateral neuralgiform headache (SUN), which only occupies cluster 17. Aside from that, cluster headaches, hemicrania continua, as well as their associated ICHD3 diagnoses probable cluster and probable hemicrania continua, occupy multiple clusters concurrently. (3, 20, 21, 22, 23, 25). The differences among these clusters appear to be only secondary to their associated autonomic features as well as indomethacin responsiveness. There is also one instance where cluster headache shares a common cluster with thunderclap headache. The distribution/clustering of various TAC here is not surprising but does highlight the potential ambiguity in separating between cluster and HC. Furthermore, the finding that there are multiple cluster groups as opposed to 1 major cluster group in migraine suggests that TAC as a class is more “fractured” in its organization – at least in our perception of it – as compared to migraine. For example, migraine with aura presents itself with multiple different kinds of subtypes also – that is, aura phenomenon - and yet remain as 1 group which is unified by, as previously noted, “reversibility” and “producibility” (i.e. more than 2 episodes). This kind of unification is not as strong in the TACs which are not only separated by the kind of autonomic features – miosis, rhinorrhea, etc. – but also by their durations.

Definitive diagnoses of frequent and chronic TTH reside in cluster 1 along with new daily persistent headache (NDPH). This is a peculiar fact but is likely due to that chronic TTH and NDPH share the characteristics of being able to be “unremitting” and “> 3 months.” “Unremitting” is not part of the migraine definition, indeed the “4 hours to 72 hours” definition eliminates this possibility and therefore eliminates the possibility of a NDPH connection. We think this is a deficit of the current classification system, as NDPH has been classically separated into “migrainous” and “tension type” phenotypes.^17^ Furthermore, we believe it is a clinically self-evident fact that there are migraine sufferers who have unremitting headaches. Therefore, we believe that allowing migraine to be “unremitting” appears to be appropriate.

Other so called miscellaneous primary headaches can be organized into their own groups – cough/Valsalva headaches, exercise headaches, traction/compression headaches, cold-induced headaches, and nummular headaches. This is to be expected since these phenomena have pathognomonic “signs”. Indeed, in a study of differential diagnosis classification, these headaches also stand out as unique.^13^

Finally, primary stabbing headaches and hypnic headaches appear to share a continuum due to their clustering in multiple groups (28, 29, 30, 31, 32, 33, 35). This can likely be explained by their criteria sharing the requirement of a lack of TAC features. This is evidenced by the fact that TAC features are broken down into the lack of miosis, rhinorrhea, etc., and these are characteristics present in the various clusters in the stabbing/hypnic headache clusters.

### Vector space

Row reduction of our matrix yields 63 characteristics as basis vector once all probable headache diagnoses are removed. This implies that those 63 characteristics, in various combinations, are sufficient to describe all definitive diagnoses of primary headaches. There are two implications of this result:

Firstly, on a clinical and practical level: since those 63 characteristics can be described by inquiring about 19 characteristics of headaches, this means that one should be able to diagnose/differentiate primary headaches from each other by using 19 questions. Of note, this is not the minimum number of questions needed to diagnose/differentiate headaches from each other. Our previous paper shows that this number can be further trimmed down.^14^ However, this result improves upon our prior result since the previous result relies heavily on duration of headaches as a means of differentiation. Duration, and frequency, as discussed previously, may not accurately describe clinical encounters; the current result, relying less so on duration, may offer a more accurate tool. Of course, our study does not include secondary headaches and so as a result, we do not recommend using the 19 questions here for new patients with red flags or potential secondary causes. Furthermore, this technique does not appear to be very beneficial for identification of probable headache disorders, as the number of characteristics in RREF for probable headaches are significant. This follows with our caveat for secondary headaches: if one is unsure about the potential diagnosis, the short cut of using 19 questions should not be used.

Secondly, the 19 question and 63 characteristics yields an important theoretical concept: since the matrix can be reduced to linear combinations of 63 pivot/vectors of characteristics, this means that definite (not probable) diagnosis of headaches lives in 63-dimensional vector space. This observation allows for a theoretical framework of measuring complexity of ICHD3, or any classification paradigm for that matter, and allows for the potential of reimaging of headaches as a Euclidean mathematical phenomenon.

## Conclusion

The mathematical embodiment of headache classification as a matrix representation offers us three potential benefits: 1) the large-scale systematic investigations of relationships between characteristics of headaches and phenotypes, 2) the graphical representation and analysis of these characteristics and phenotypes, and 3) the potential improvement in clinical diagnosis of headaches both by automation and elimination of redundancy in history taking.

## Supporting information

Supplemental Material - Matrix

## Data Availability

All data produced in the present study are available upon reasonable request to the authors

## Statement of authorship

Category 1

(a) Conception and Design: Roger Cheng MD, Pengfei Zhang, MD
(b) Acquisition of Data: Pengfei Zhang, MD
(c) Analysis and Interpretation of Data: Roger Cheng MD, Pengfei Zhang, MD

Category 2

(a) Drafting the Manuscript: Roger Cheng MD, Pengfei Zhang, MD
(b) Revising It for Intellectual Content: Roger Cheng MD, Pengfei Zhang, MD

Category 3

(a) Final Approval of the Completed Manuscript: Roger Cheng MD, Pengfei Zhang, MD

## Acknowledgements

None

## Abbreviations

ICHD3: International Classification of Headache Disorders, 3^rd^ Edition
TAC: trigeminal autonomic cephalalgia
RREF: row reduction eschelon form
TTH: tension-type headache
HC: hemicrania continua
PH: paroxysmal hemicrania
NDPH: new daily persistent headache

## References

1. International Headache Society. International Classification of Headache Disorders. Cephalalgia. (2018) 38:1–211. doi: 10.1177/0333102417738202

2. Costabile R, Catalano G, Cuteri B, Morelli MC, Leone N, and Manna M. A logic-based decision support system for the diagnosis of headache disorders according to the ICHD-3 international classification. Theory Prac Log Prog 2020;20(6):864–879.

3. Zhang P. Which headache disorders can be diagnosed concurrently? An analysis of ICHD3 criteria using prime encoding system. Front Neurol. 2023;14:1221209

4. Weisstein E. “Bipartite Graph.” From MathWorld--A Wolfram Resource. https://mathworld.wolfram.com/BipartiteGraph.html (Accessed December 6, 2025)

5. Domagalski R, Neal ZP, Sagan B. Backbone: An R package for extracting the backbone of bipartite projections. PLoS One. 2021 Jan 6;16(1):e0244363. doi: 10.1371/journal.pone.0244363. PMID: 33406145; PMCID: PMC7787471.

6. Ma E. Network Analysis Made Simple. https://github.com/ericmjl/Network-Analysis-Made-Simple. (Accessed December 6, 2025)

7. Hoffman M. Methods for Network Analysis. https://bookdown.org/markhoff/social_network_analysis/. (Accessed December 6, 2025)

8. Lee D, Seung HS. Algorithms for nonnegative matrix factorization. In: Leen T, Dietterich T, Tresp V, editors. Advances in neural information processing systems. Vol. 13. Cambridge, MA: MIT Press; 2000.

9. van Dongen, Stijn, Graph clustering via a discrete uncoupling process, Siam Journal on Matrix Analysis and Applications 30–1, p121-141, 2008. (10.1137/040608635).

10. Allard G. Markov Clsutering. https://markov-clustering.readthedocs.io/en/latest/readme.html. (Accessed December 6, 2025)

11. Malliaros, Fragkiskos D., and Michalis Vazirgiannis. “Clustering and community detection in directed networks: A survey.” Physics Reports 533.4 (2013): 95–142

12. Shannon P, Markiel A, Ozier O, Baliga NS, Wang JT, Ramage D, Amin N, Schwikowski B, Ideker T. Cytoscape: a software environment for integrated models of biomolecular interaction networks. Genome Res, 13:11 (2498-504). 2003 Nov. PubMed ID: 14597658.

13. Zhang, P. The Structure and Organizations of ICHD-3 Differential Diagnoses through DiffNet: A Pilot Study. Diagnostics 2022, 12, 2589. 10.3390/diagnostics12112589

14. Zheng P, Cheng R. Diagnosis Through Differentiation: A Pilot Study on Improving the Diagnostic Efficiency of Primary Headaches in ICHD3. Frontiers in Neurology, Accepted December 5, 2025.

15. Lay DC. Linear Algebra and Its Applications. Pearson Education; 2003.

16. Zhang P, Berk T. Network analysis of headache diagnoses using international classification of headache disorders, 3rd edition. Front Neurol. 2025 Jan 30;16:1526037. doi: 10.3389/fneur.2025.1526037. PMID: 39949792; PMCID: PMC11824271.

17. Robbins MS, Crystal SC. New daily-persistent headache versus tension-type headache. Curr Pain Headache Rep. 2010 Dec;14(6):431–5. doi: 10.1007/s11916-010-0145-3. PMID: 20865354.

18. Abstracts from the 18 th European Headache Congress (EHC) : Rotterdam, The Netherlands. 4-7 December 2024. J Headache Pain. 2025 Jun 23;26(Suppl 2):138. doi: 10.1186/s10194-025-02062-8. PMID: 40545525; PMCID: PMC12183827.

